# Diagnostic Accuracy of WHO IMCI Visual Inspection for Neonatal Jaundice: A Prospective Multi-Country Cohort

**DOI:** 10.64898/2026.09.16.26362263

**Authors:** Amna Khan, Alyssa Shapiro, Nida Salman Yazdani, Sebin George Abrahim, Victor Akelo, Kwaku Poku Asante, Florence Aweyo, Anne George Cherian, Munita Jat, Fyezah Jehan, Indhumathi K, Kevin Kasadhe, Margaret P. Kasaro, Anne CC Lee, Azqa Mazhar, Sarmila Mazumder, Christopher N. Mores, Wilbroad Mutale, Humphrey Mwape, Sam Newton, Muhammad Imran Nisar, Linda Ogala, Nancy Ogola, Neeraj Sharma, M. Bridget Spelke, Jasmine Sugirtha, Yipeng Wei, Wen-Chien Yang, Zahra Hoodbhoy, Qing Pan, Emily R. Smith

## Abstract

**BACKGROUND:** Adverse consequences of neonatal hyperbilirubinemia disproportionately affect babies in low- and middle-income countries. Laboratory measurement of total serum bilirubin is limited in low-resource settings, necessitating simpler alternatives. Our study aimed to evaluate the diagnostic accuracy of visual inspection of jaundice via the World Health Organization Integrated Management of Childhood Illnesses algorithm compared to transcutaneous bilirubinometry for the identification of hyperbilirubinemia in community settings.

**METHODS:** Hyperbilirubinemia was assessed by visual inspection for jaundice and transcutaneous bilirubinometry at 0-72 hours, 3-5 days, and 5-14 days postnatal in the PRISMA prospective cohort study in six sites in Asia and Africa. We assessed sensitivity, specificity, and positive and negative percent agreement.

**RESULTS:** We analyzed data from 14,331 infants, including 1,852 preterm infants. 15% had hyperbilirubinemia, as defined by transcutaneous bilirubin values and the NICE nomograms, at 3-5 days postnatal. Visual inspection had a sensitivity of 16%, specificity of 96%, positive percent agreement of 56%, and negative percent agreement of 86% at 3-5 days postnatal age, pooled across all sites and gestational ages, versus transcutaneous bilirubin defined by NICE nomograms.

**CONCLUSION:** The poor sensitivity of visual inspection underscores an urgent need for accurate, affordable screening tools to improve screening for hyperbilirubinemia.

**Impact Statement:**

- We found poor sensitivity of visual inspection, identifying fewer than one in five neonatal jaundice cases, suggesting much worse diagnostic accuracy in this community-based context across five low- and middle-income countries (LMICs) as compared to the previous studies conducted in health facilities.
- Among preterm infants, sensitivity was modestly higher than in term infants but remained clinically inadequate, reinforcing that visual inspection alone is insufficient for this high-risk group.
- Despite standardized training and biannual refresher sessions, our study suggests that training for health care workers alone is unlikely to make visual inspection a reliable case-detection tool.
- Our findings suggest that the WHO should re-evaluate the IMCI recommendations to include tools with higher diagnostic accuracy to identify neonatal jaundice.
- Community-level jaundice screening should move beyond visual inspection alone towards validated, low-cost, objective tools; icterometers and smartphone apps with high diagnostic accuracy are promising alternatives.

## Introduction

Neonatal jaundice affects approximately 50% of term and 80% of preterm infants globally,^1^ with an estimated 1.1 million infants developing severe neonatal jaundice requiring medical interventions.^2^ Neonatal jaundice ranks seventh among causes of neonatal death globally,^3^ with higher proportions of mortality in low- and middle-income countries (1.19 per 1000 livebirths) as compared to high-income countries (0.01 per 1000 livebirths).^4^ A recent systematic review suggested that 8-31% of hospitalized jaundiced infants develop severe neonatal jaundice, a high-risk condition indicated by the presence of acute bilirubin encephalopathy and need for exchange blood transfusion; the estimated global burden of severe neonatal jaundice is 67 and 25 cases per 1000 live births in Sub-Saharan Africa and South Asia, respectively.^5^ This disparity can be attributed to widespread challenges in healthcare access and delivery systems.^6^ Therefore, early detection is especially important in low- and middle-income countries to prevent complications of neonatal jaundice and reduce hospital admissions, where treatment remains limited.

The gold standard for neonatal jaundice diagnosis is total serum bilirubin measurement. However, serum bilirubin measurement is often inaccessible in low- and middle-income countries due to infrastructure limitations including cost and electricity.^7^ Serum bilirubin measurements require invasive blood draws; thus, parents and providers prefer noninvasive methods when possible.^8,9^ Transcutaneous bilirubinometry, assessed by shining light on the skin and measuring reflected or transmitted light, provides a practical, quick, and pain-free alternative. Studies report that transcutaneous bilirubinometry has a strong correlation with serum bilirubin (r = 0.82, 95% CI 0.78-0.85), supporting its use as a reliable, non-invasive option.^10,11^ However, these results need to be interpreted with caution, as the reliability of transcutaneous bilirubinometry for assessing preterm infants and infants with dark skin tones has limitations and requires robust studies to establish its effectiveness and reliability for this population.^12^ In addition, the high cost of transcutaneous devices – around USD 2600–8200 for a single unit – makes it inaccessible for resource-limited settings.^13^ Currently, clinical practice guidelines recommend transcutaneous bilirubinometry as an accurate, reliable screening tool only, not as a substitute for serum bilirubin, and that transcutaneous values indicating that a newborn is at risk of hyperbilirubinemia should be followed up with a serum bilirubin measurement;^14,15^ however, evidence from LMICs suggests that transcutaneous bilirubinometry is often used as a substitute for serum bilirubin measurement when this tool is unavailable.^16^

Visual inspection remains the frontline approach for screening for newborn jaundice in primary health care or community settings where serum and/or transcutaneous testing is unavailable. It follows established frameworks such as the Kramer scale, which uses skin blanching to assess jaundice progression in an infant from head to toe and estimate bilirubin levels.^17,18^ The widely used World Health Organization Integrated Management of Childhood Illness (IMCI) strategy provides community health workers with standardized approaches for the identification and management of common childhood illnesses, including identification of jaundice via visual inspection.^19^ It categorizes jaundice into three levels of severity – not jaundiced, jaundiced, and severely jaundiced, defined as jaundice of the palms/soles at any time or anywhere on the body at < 24 hours of life. However, existing literature on visual inspection reports inconsistent performance; despite its ease and effectiveness, visual inspection is subjective, with accuracy influenced by ambient lighting, skin pigmentation, and observer experience.^17,20–22^ Given these limitations, more evidence is needed to expand our understanding of its utility as a screening tool, particularly in darker skin tones.^22^

There are other evidence gaps on this topic. Evidence on visual inspection in low- and middle-income countries is limited and remains largely hospital-based, while most first-line newborn care happens in primary care and community settings, with a wide variability in reported sensitivity and specificity for identifying hyperbilirubinemia.^23^ Also, community-based evaluations of visual inspection are scarce, and preterm infants are mostly underrepresented. Direct comparisons between visual inspection and transcutaneous bilirubinometry remain unexplored. To address this gap, our primary objective was to assess and compare the diagnostic accuracy of the current IMCI jaundice algorithm compared to transcutaneous bilirubinometry for identifying term and preterm newborns for hyperbilirubinemia in community settings in six study sites in Asia and Africa.

## Methods

### Study design and setting

This study leverages data collected in the Pregnancy Risk, Infant Surveillance, and Measurement Alliance (PRISMA) study (Clinicaltrials.gov trial registration number: NCT05904145). The PRISMA study is a prospective open cohort study at six research sites across five countries (Kintampo, Ghana; Kisumu, Kenya; Lusaka, Zambia; Karachi, Pakistan; Vellore, south India; and Hodal: north India).^24^ Across all sites, assessments were performed in community settings (at primary health care centers and participants’ homes) rather than in tertiary hospitals. Pregnant women who lived in the catchment areas were enrolled consecutively before a gestational age of 20 weeks if they met the minimum required age (Ghana, Zambia, and Pakistan: 15 years; Kenya and India: 18 years) and provided written informed consent. The present study utilized data collected between September 2022 and December 2025 and included all liveborn singletons, twins, or triplets with at least one postnatal assessment that included both visual inspection and transcutaneous bilirubin measurement. If infants were enrolled but never assessed (e.g., early death, loss to follow-up before first visit, or hospitalization), they were excluded from analysis.

### Study procedures and data collection

As part of the PRISMA study, all newborns received standardized postnatal care visits conducted by research personnel. Jaundice was systematically evaluated during three scheduled visits: within 72 hours after birth, 3–5 days postnatal, and 5–14 days postnatal. Assessments included both noninvasive transcutaneous bilirubin (TCB) measurement using the BiliCare® System (Model: 81000200) and visual inspection for jaundice using the World Health Organization (WHO) Integrated Management of Childhood Illness (IMCI) method, performed per standardized protocols. The BiliCare transcutaneous device has been previously validated against serum bilirubin in several studies, with sensitivity and specificity reported as 78-85% and 79-94%, respectively.^12,25^ To ensure measurement consistency, we used harmonized clinical protocols and conducted regular refresher training across study sites. Certified assessors evaluated infants unclothed under well-lit conditions, examining three body regions: 1) head (face, gums, sclera), 2) trunk (chest, abdomen), and 3) distal extremities (palms, soles).^19^ Based on Integrated Management of Childhood Illness guidelines, jaundice severity was classified into one of the three categories: “Not Jaundiced” if no yellow discoloration was observed, “Jaundiced” if they had yellow eyes and skin, and “Severely Jaundiced” if yellow discoloration appeared anywhere on the body in the first 24 hours of life or had spread to palms and soles at any time point.^26^ Following visual inspection, non-invasive transcutaneous bilirubin levels were measured once using the BiliCare® device.^27^ Results were instantly displayed on the device and recorded in electronic case report forms. To maintain data integrity, the device underwent routine daily calibration checks, ensuring standardized measurements across all sites, and data was reviewed biweekly to identify potential outliers or errors. Calibration checks were done by placing a calibration check tip, provided by the manufacturer, over the Bilicare sensor and taking a measurement; the device would then display a message that the calibration check was successful. Research staff performed transcutaneous bilirubin measurements after visual inspection, so they knew the visual inspection result prior to the transcutaneous result but not vice versa.

For both visual inspection and transcutaneous bilirubin measurements, if an infant was measured twice in the same time period, only the first data point was included. Additionally, if any measurements occurred outside of the period of 0 to ≤ 14 days postnatal, this data was excluded from analysis. Infants were referred for further clinical evaluation per local treatment guidelines based on results of visual inspection.

### Outcome definitions

Visual inspection served as the index test and TCB as the reference standard for clinically significant hyperbilirubinemia; serum bilirubin, the true gold standard, was not measured as part of the study protocol. For both the index test and the reference standard, we classified an infant as “Positive” or “Negative” for jaundice/hyperbilirubinemia at each timepoint, then assessed how well the two agreed.^19^

Transcutaneous bilirubinometry was used as the reference standard to identify clinically significant hyperbilirubinemia. The three methods were applied independently, and each infant was classified as positive/negative for clinically significant hyperbilirubinemia under each method. 1) *Static absolute value cutoff method:* transcutaneous bilirubin values of 15 mg/dL or greater for term babies (gestational age at birth of ≥ 37 weeks) and ≥ 10 mg/dL for preterm babies were defined as “Positive.” 15 mg/dL is recommended for term newborns by the AAP guidelines; while guidelines for preterm infants do not have a clear consensus, we selected 10 mg/dL as a middle ground between the values of 5-14 mg/dL for gestational ages of 28-35 weeks, respectively, suggested by Maisels et al in their supplement to the 2004 version of the AAP guidelines ^28^. 2) *NICE Nomogram Method:* This method applies nomograms developed by the National Institute of Health and Care Excellence (NICE) in the UK, which provides total serum bilirubin (TSB) values at which phototherapy is recommended based on the gestational age, neonatal age, and risk factors of the newborn.^14^ We applied a decision rule of −3 mg/dL below the TSB threshold, a common practice when using transcutaneous instead of serum bilirubin values under the NICE or American Academy of Pediatrics (AAP) guidelines.^29,30^ This means that if a transcutaneous value is [“serum bilirubin threshold at which phototherapy is recommended” – 3] mg/dL or higher, we classified this newborn as “Positive”. 3) *AAP Nomogram Method:* We classified a newborn as “Positive” using the nomograms in the American Academy of Pediatrics (AAP) guidelines.^15^ If a newborn had a GA at birth of less than 35 weeks, they were excluded from analysis by the AAP Nomogram Method only. If the newborn had a diagnosis of birth asphyxia (defined as clinician-reported failure to breathe spontaneously in the first minute after delivery or breathing assistance was required) or sepsis as suspected (by a clinician) or proven (with culture)), we classified them using the AAP nomograms for infants with hyperbilirubinemia neurotoxicity risk factors, which use slightly lower thresholds.^15^

### Data Analysis

First, we excluded data from any postnatal visit where either the index or reference test was missing or not conducted on the same visit day. We then presented the descriptive characteristics of newborns and their mothers, along with a summary of jaundice prevalence by transcutaneous bilirubin and visual inspection assessment. Because transcutaneous bilirubinometry is an imperfect proxy for the serum bilirubin gold standard, we report the agreement between the two tests as our primary outcomes: positive percent agreement (PPA; the proportion of transcutaneous-positive infants that visual inspection also classified as positive) and negative percent agreement (NPA; the proportion of transcutaneous-negative infants that visual inspection also classified as negative) rather than positive predictive value or negative predictive value. Then, we also reported the sensitivity and specificity of visual inspection vs. transcutaneous bilirubinometry for diagnosis of clinically significant hyperbilirubinemia.

## Results

Results are reported according to the STARD 2015 guidelines for studies of diagnostic accuracy; a completed STARD checklist can be found in **Table S1.**^31^

### Participants characteristics

A total of 17,821 pregnant individuals (18,635 pregnancies) were enrolled in PRISMA between September 2022 and December 2025, and 14,945 infants were included in this analysis, generating 41,152 unique data points **(Figure 1)**. After excluding cases with missing or invalid values for neonatal age, visual inspection results, age, or TCB results, and cases with multiple readings in the same time frame, a total of 30,624 unique study visits from 14,331 infants (of 13,816 mothers) were considered for the main analysis. In the included sample, the majority of infants were term births (87%), with some meaningful site-specific differences, such as higher percentages of preterm births in Pakistan **(Table 1)**. No adverse events were reported from visual inspection or transcutaneous bilirubin measurement. 4.3 percent of infants were identified as ‘Positive’ for jaundice by visual inspection in the 3-5 days postnatal time period; 15% had hyperbilirubinemia, as defined by the NICE nomograms, in the 3-5 days postnatal time period **(Table S2)**.

**Figure 1.**
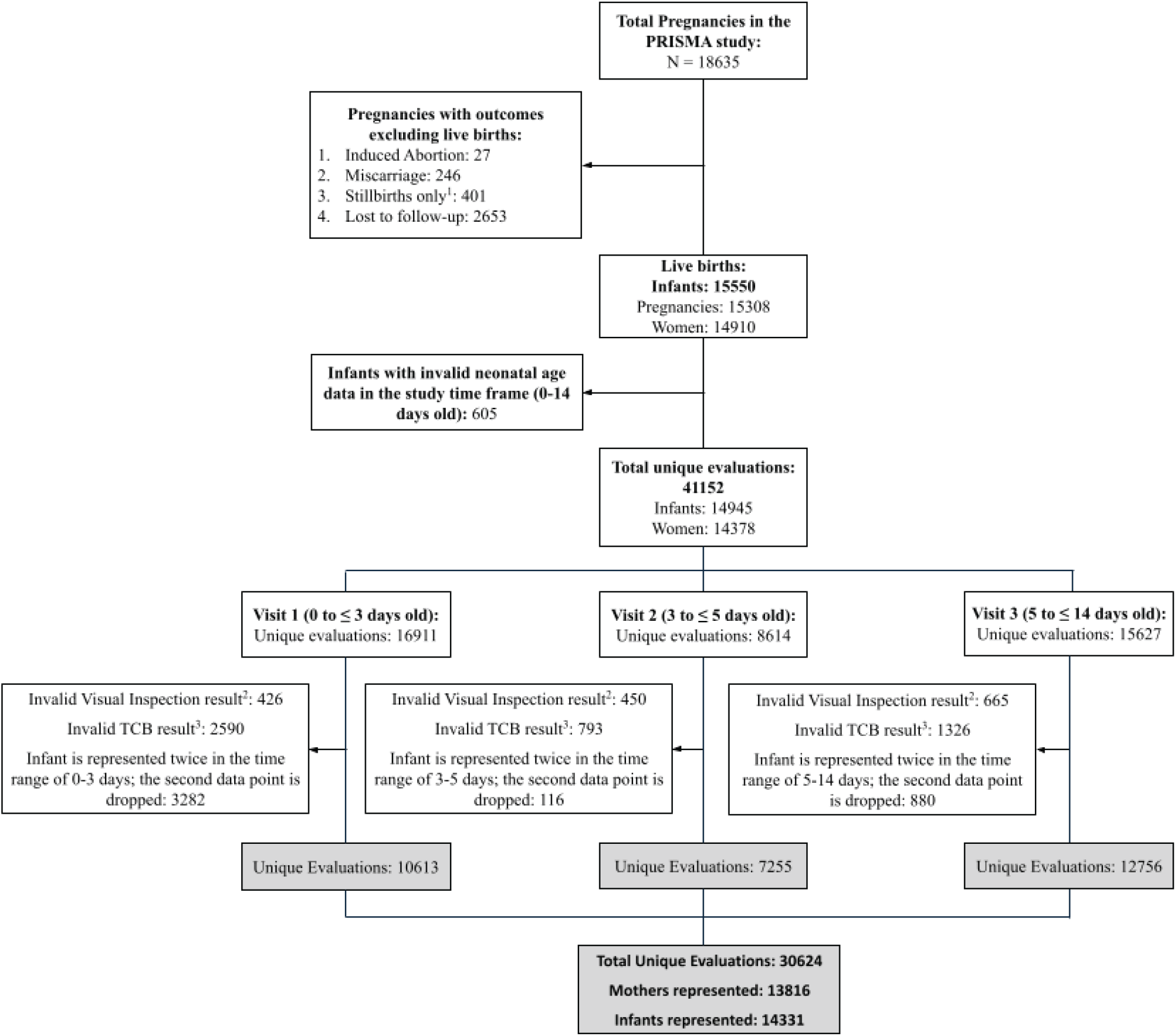
Flow diagram of pregnant participants and infants selected for analysis. ^1^In 12 cases, a woman was pregnant with twins but the outcome resulted in one live birth and one stillbirth. These instances were not counted here in the n=401 number but were instead counted in the subsequent box (live births) as 1 pregnancy and 1 infant. ^2^Valid Visual Inspection result: excludes answers marked as NA or ‘Don‘t know’ ^3^Valid TCB: excludes answers marked as NA and outside the range of the device (0-20 mg/dL)

**Table 1.** Maternal and neonatal characteristics.

| Characteristic | Overall | North India | South India | Pakistan | Ghana | Kenya | Zambia |
| --- | --- | --- | --- | --- | --- | --- | --- |
| **Maternal Factors** |  |  |  |  |  |  |  |
| N = | 13,816 | 1,789 | 1,996 | 4,461 | 1,990 | 2,213 | 1,367 |
| Age | 26.5 ± 5.6 | 23.8 ± 3.7 | 25.2 ± 4.4 | 27.0 ± 5.9 | 27.8 ± 6.3 | 27.0 ± 5.4 | 27.9 ± 6.0 |
| Education |  |  |  |  |  |  |  |
| No education | 3,615 (26%) | 202 (11%) | 9 (0.5%) | 2,601 (58%) | 780 (39%) | 10 (0.5%) | 13 (1.0%) |
| School < 10 years | 3,950 (29%) | 468 (26%) | 139 (7.0%) | 1,226 (27%) | 758 (38%) | 653 (30%) | 706 (52%) |
| School > 10 years | 6,239 (45%) | 1,119 (63%) | 1,848 (93%) | 634 (14%) | 450 (23%) | 1,550 (70%) | 638 (47%) |
| **Labour & Delivery** |  |  |  |  |  |  |  |
| N = | 14,331 | 1,798 | 2,053 | 4,780 | 2,036 | 2,275 | 1,389 |
| Gestational Age at Delivery ≥ 37 weeks (Term) | 12,479 (87%) | 1,551 (86%) | 1,896 (92%) | 3,786 (79%) | 1,922 (94%) | 2,090 (92%) | 1,234 (89%) |
| Place of Birth |  |  |  |  |  |  |  |
| Health care facility | 13,102 (93%) | 1,741 (97%) | 2,030 (100%) | 4,017 (85%) | 1,798 (90%) | 2,176 (97%) | 1,340 (98%) |
| Homebirth | 1,034 (7.3%) | 51 (2.8%) | 0 (0%) | 704 (15%) | 195 (9.8%) | 61 (2.7%) | 23 (1.7%) |
| Mode of Delivery |  |  |  |  |  |  |  |
| Spontaneous Vaginal Delivery | 10,654 (74%) | 1,353 (75%) | 1,518 (74%) | 3,299 (69%) | 1,728 (85%) | 1,816 (80%) | 940 (68%) |
| C-section | 3,673 (26%) | 445 (25%) | 535 (26%) | 1,481 (31%) | 308 (15%) | 457 (20%) | 447 (32%) |
| **Infant Factors** |  |  |  |  |  |  |  |
| N = | 14,331 | 1,798 | 2,053 | 4,780 | 2,036 | 2,275 | 1,389 |
| Singleton Births | 13,946 (99%) | 1,784 (100%) | 2,008 (99%) | 4,661 (99%) | 1,946 (98%) | 2,196 (98%) | 1,351 (98%) |
| Male Sex | 7,334 (51%) | 955 (53%) | 1,087 (53%) | 2,464 (52%) | 1,002 (49%) | 1,112 (49%) | 714 (52%) |
| Birth Centiles |  |  |  |  |  |  |  |
| Small for Gestational Age (SGA) | 4,125 (29%) | 602 (33%) | 448 (22%) | 1,638 (34%) | 621 (31%) | 431 (19%) | 385 (28%) |
| Appropriate for Gestational Age (AGA) | 9,957 (69%) | 1,179 (66%) | 1,569 (76%) | 3,073 (64%) | 1,380 (68%) | 1,783 (78%) | 973 (70%) |
| Large for Gestational Age (LGA) | 249 (1.7%) | 17 (0.9%) | 36 (1.8%) | 69 (1.4%) | 35 (1.7%) | 61 (2.7%) | 31 (2.2%) |
| Birth Weight (g) | 2,787 ± 668 | 2,752 ± 436 | 2,923 ± 440 | 2,565 ± 812 | 2,889 ± 611 | 3,018 ± 543 | 2,874 ± 678 |
| Low birth weight (<2500 g) | 2,646 (18%) | 423 (24%) | 308 (15%) | 1,178 (25%) | 270 (13%) | 249 (11%) | 218 (16%) |
| Birth Asphyxia<sup>1</sup> | 1,097 (7.7%) | 158 (8.8%) | 131 (6.4%) | 411 (8.6%) | 202 (9.9%) | 99 (4.4%) | 96 (6.9%) |
| Neonatal Sepsis<sup>2</sup> | 176 (1.2%) | 5 (0.3%) | 19 (0.9%) | 24 (0.5%) | 9 (0.4%) | 35 (1.5%) | 84 (6.0%) |

Categorical variables are displayed as number and percentage of the total; Continuous variables are displayed as mean and standard deviation.
<sup>1</sup>Defined as when a clinician reports failure to breathe spontaneously in the first minute after delivery or breathing assistance was required.
<sup>2</sup>Defined as inflammatory response and organ dysfunction following presence of a severe infection from delivery to 28 days as suspected (by a clinician) or proven (with culture).

### Diagnostic performance of visual inspection

The diagnostic performance of visual inspection against transcutaneous bilirubin assessments based on three methods are shown in **Figure 2** and **Figure S1**. In **Figure 2A**, a static cutoff method (transcutaneous bilirubin walue ≥ 15 in Derm babies or ≥ 10 in preDerm babies) was used. Term and preterm neonates are shown separately in **Figures S1A and S1B,** respectively. Visual inspection had poor sensitivity overall; it was ≤ 32% for all sites and at all timepoints, with some heterogeneity across sites. Specificity was ≥ 97%, and negative percent agreement was ≥ 82% for all sites at all time points. The static, lower transcutaneous bilirubin cutoff (≥ 10) for preterm infants **(Figure S1B)** resulted in a higher positive percent agreement but lower sensitivity and negative percent agreement, compared to using the same value (≥15) for preterm and term infants **(Figure S1C).** Thus, specifying different cutoffs for preterm infants and term infants improves the positive percent agreement – with minimal effect on sensitivity and negative percent agreement – compared to using the same cutoff of ≥ 15 for all infants **(Figure S1D).**

**Figure 2.**
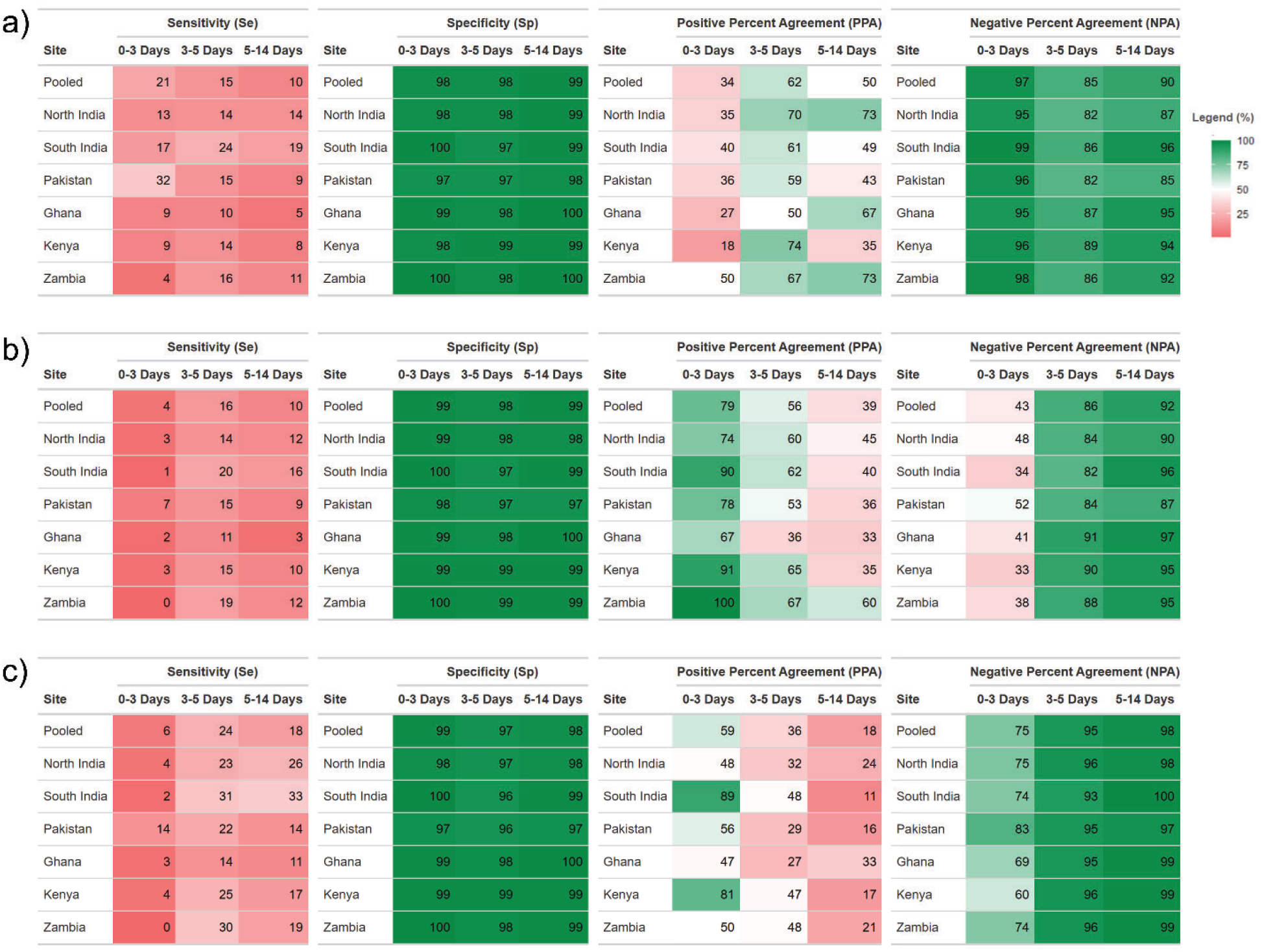
Diagnostic performance of visual inspection against transcutaneous bilirubin, by a) Static Cutoff Method: Transcutaneous bilirubin ≥ 15 for term infants and ≥ 10 for preterm infants, b) NICE Nomogram Method: Transcutaneous bilirubin ≥ NICE cutoffs, for all babies, c) AAP Nomogram Method: Transcutaneous bilirubin ≥ AAP cutoffs, for all babies with gestational age ≥ 35 weeks

In the diagnostic performance evaluation against transcutaneous bilirubin defined by the NICE nomogram (**Figure 2B),** the sensitivity and negative percent agreement of visual inspection compared to transcutaneous bilirubin defined by the NICE Nomogram method are significantly lower than when comparing to transcutaneous bilirubin defined by the static cutoff method **(Figure 2A)** for the first three days of life. However, similar patterns are seen for specificity and negative percent agreement in the 0–3 days age window for both the NICE Nomogram method and the Static Cutoff methods. Also, the diagnostic performance of visual inspection compared to transcutaneous bilirubin–based diagnosis using the AAP thresholds for clinically indicated hyperbilirubinemia was very similar to that using the NICE nomogram method (**Figure 2C**).

## Discussion

This is among the first and largest studies to test the WHO IMCI visual inspection method against transcutaneous bilirubin in community settings to identify the need for referral for clinically significant hyperbilirubinemia. Across the five countries, visual inspection failed to identify most infants who met transcutaneous bilirubin thresholds for neonatal jaundice referral, correctly detecting fewer than one in five true cases. For a frontline screening tool, this represents a clinically unacceptable failure rate; the infants missed are precisely those at risk of progressing to severe jaundice and its irreversible neurological complications. Our findings suggest that the current World Health Organization Integrated Management of Childhood Illnesses method for jaundice identification should be updated in order to improve its ability to identify at-risk cases.

Similar to previous evidence, visual inspection demonstrated poor sensitivity and positive percent agreement at all sites, though with wide variability across sites. Compared with earlier studies reporting varied sensitivity and more modest negative predictive values, our findings suggest that the primary limitation of visual inspection lies in the under-identification of true cases rather than over-classification of infants as “Positive” for jaundice when they are healthy, a pattern that persisted under both NICE and AAP transcutaneous bilirubin thresholds.^17,20,21,32–34^ Negative percent agreement was consistently high (82-100%) across all sites. Where transcutaneous and serum bilirubin measurements are unavailable, a visual inspection result of no or mild yellow discoloration may offer some reassurance that an infant does not require urgent referral for jaundice. This reassurance, however, operates only at the population level and does not offset the danger at the individual level, since visual inspection still misses many true cases. Our study addressed an important evidence gap on the topic: there is a paucity of evidence on the diagnostic accuracy of visual inspection from community settings, where many newborns in LMICs are first assessed. To our knowledge, Hatzenbuehler et al. in Pakistan was the only study that similarly evaluated visual inspection (defined using the Kramer method) in community settings.^35^ Data was collected by three primary healthcare workers evaluating 143 newborns, and they found a sensitivity of 83.3% and specificity of 50.5%. Our sensitivity was lower and specificity higher in Pakistan (sensitivity=15% and specificity=97% among newborns 3-5 days old), but our Pakistan site included 20 healthcare workers and 4,780 newborns. The wider pool of observers and larger sample in our study captures the inter-observer variability that would be expected in routine community health programs but is masked in small, tightly controlled studies.

Most prior studies on the diagnostic accuracy of visual inspection were done in hospital settings, where level of health provider, lighting, and hyperbilirubinemia risk differ. Hospital-based studies evaluating Kramer’s method of visual inspection^18^ by Keren et al in the US (NPV=84%) and Aprillia et al in Indonesia (NPV=89%) support the use of visual inspection for ruling out jaundice in healthy newborns.^17,36^ Conversely, hospital-based studies evaluating Kramer’s method by Dionis et al in Tanzania and Sampurna et al in Indonesia reached opposite conclusions, finding that visual inspection cannot be used to rule out jaundice in healthy newborns, with negative predictive values of 63% and 81%, respectively.^22,34^ Although we did not find any studies evaluating visual inspection by the IMCI method in community settings, three studies evaluated visual inspection by the IMCI method in hospital settings;^37–39^ Kanodia et al and Thummakomma et al found higher sensitivity values than the current study (100% and 61.35%, respectively), and Kaur et al did not do a direct calculation of diagnostic accuracy. However, we expect that in tertiary hospital settings, jaundice prevalence and severity are likely to be higher than in community settings, fewer staff members take measurements, and lighting and clinical conditions are controlled. These factors collectively inflate diagnostic accuracy estimates relative to what would be expected in routine community-based screening. Research staff in this study evaluated newborns for jaundice in natural light settings, and the importance of good lighting conditions was emphasized in standard operating procedures and trainings, but the quality of this lighting could still vary depending on air pollution, time of day, and the setting (home, primary health center, etc.) where measurements took place.

Second, we found it difficult to directly compare our results to the larger literature due to the heterogeneity in how visual inspection positivity is defined. Some studies defined positive cases of jaundice using different Kramer score thresholds, and others defined a positive case as any visible jaundice. In contrast, the WHO-IMCI definition of “Severely Jaundiced” corresponds to only a 5 on the Kramer scale, a more conservative threshold which likely contributed to lower sensitivity in our study. Dionis et al. classified all newborns with a Kramer score between 1 to 5 as positive and reported a sensitivity of 71% and a positive predictive value of 90%.^22^ Darmstadt et al evaluated their own definitions of yellow discoloration and endorsed the definition ‘any jaundice of the distal extremities or deep jaundice of the trunk or head’.^23^ Other definitions of Positive for neonatal jaundice included Kramer scores of 1-5,^36,40^ 3-5,^33,34^ and 4-5.^35^ Thus, while our sensitivity numbers are lower than those found in the literature, they must be interpreted with consideration for these differences in the definition of “Positive” for jaundice by the index test (visual inspection). Further research should evaluate whether modifications to the WHO IMCI visual inspection criteria, such as adopting a lower Kramer threshold or incorporating structured anatomical scoring, could improve sensitivity while retaining feasibility for community health workers.

Preterm infants are at higher risk for neonatal jaundice and its complications,^1^ and our study is one of the first to assess the diagnostic accuracy of visual inspection for a large (n=1,852) sample size of preterm infants in community settings. Our study found that sensitivity of visual inspection compared to transcutaneous bilirubin values of ≥ 15 mg/dL was slightly better but comparable for preterm infants 3-5 days old vs. term infants 3-5 days old, while specificity and negative predictive value were generally comparable. These findings support that visual inspection has a better but still suboptimal ability to detect jaundice in at-risk preterm infants. Szabo et al evaluated a small sample size of 69 preterm infants and also concluded that the Kramer visual inspection method had poor accuracy for detecting significant neonatal jaundice in preterm infants.^41^

Our findings highlight the urgent need for alternative low-cost tools for accurate detection of jaundice in LMICs. Icterometers, such as the Bili-ruler and Bili-strip, have demonstrated high sensitivity in recent pilot studies conducted in LMICs.^42–44^ The Bili-ruler costs just $40 USD, has CE mark, and was also evaluated in the PRISMA study;^45^ results will be published soon. Development of low-cost smartphone apps has also progressed rapidly in recent years; the Picterus app, for example, also has CE mark and was recently evaluated in Mexico, Nepal, and the Phillipines, where it was shown to have a Pearson’s correlation coefficient of r=0.76 compared to total serum bilirubin.^46^ Finally, portable serum bilirubin measurement tools, such as BiliDx and BiliStick, both have CE marks and could provide accurate measurement of serum bilirubin in community settings, though the cost of these tools - while much more affordable than laboratory serum bilirubin - could be unaffordable in many settings.^47–49^

Our study has several notable strengths. First, it is one of the largest community-based evaluations of neonatal jaundice screening in low- and middle-income countries to date, across six community settings in five LMICs, and it includes a large sample size of preterm infants. Nearly all prior diagnostic accuracy studies of visual inspection were derived from hospital settings, where jaundice prevalence, observer expertise, severity and cases differ from the primary care and household settings. Second, as the WHO Integrated Management of Childhood Illnesses guidelines are widely adopted in low-resource settings, it is important to examine the ability of this technique to identify neonatal jaundice that requires follow-up care. Our study benefits from the inclusion of many research staff trained in WHO IMCI visual inspection techniques, which better reflects LMIC settings under which visual inspection is performed in community health programs. Additionally, we incorporated standardized training, including biannual refresher training, on the IMCI method of jaundice identification, which strengthens the internal validity of comparisons across multiple research sites. However, there are a few limitations in our study. First, we used non-invasive transcutaneous bilirubinometry as our reference standard because it is more often used in community settings; however, serum bilirubin remains the true gold standard tool for bilirubin evaluation, and using transcutaneous bilirubinometry introduces imperfect reference standard bias. While transcutaneous bilirubinometry may be less reliable in some populations, such as dark-skinned and preterm newborns, recent reviews suggest a high correlation between transcutaneous and serum bilirubin measurement in preterm infants.^50,51^ Additionally, while the BiliCare brand of transcutaneous bilirubinometer device has been validated in several low- and middle-income countries compared to serum bilirubin assessment,^11,12,52,53^ data evaluating the BiliCare in dark-skinned African populations has yet to be published, which is a critical gap given that both visual inspection and transcutaneous bilirubinometry accuracy may be affected by skin pigmentation.^54^ However, we have found that even in Kenyan, Zambian and Ghanaian newborns in this study, there are very few infants with dark skin based on objective measures and likely due to the low melanin in skin in the newborn period. Thus, the true effect of skin pigmentation on BiliCare accuracy in this neonatal population may be uncertain. Also, inter-individual variation remained high, even with high-quality, standardized training and biannual refresher sessions, and this finding suggests that training alone is unlikely to make visual inspection by the IMCI method a reliable case-detection tool. This is consistent with the Darmstadt et al study,^23^ which evaluated several definitions of newborn jaundice by visual inspection in six hospitals and likewise found a high variation between the study sites (for example, the definition of “deep jaundice of the trunk or head” had a sensitivity varying from 55-98% among physicians at the six sites). Finally, the NICE guidelines (UK) and AAP guidelines (USA) may be limited in their applicability to low- and middle-income countries. The thresholds provided are based on a combination of expert consensus – derived by experts who are mostly from high-income settings – and evidence collected in high-resource hospitals, particularly these ^55^.

A review by Okwundu and Saini concludes that visual inspection is the least reliable of bilirubin estimation methods and should not be used at all to assess newborns for jaundice.^32^ Taken together, our findings of low sensitivity and positive percent agreement, along with the mixed negative predictive value findings in the literature,^17,22,34–36^ indicate that visual inspection via the WHO IMCI technique should not be used as a stand-alone screening tool, reinforcing current guidance.^14,15^ that visual assessment is an initial step to screen out cases rather than a substitute for transcutaneous or serum testing, where available. Building on these findings, we propose three concrete next steps. First, community-level jaundice screening should move beyond visual inspection alone towards validated, low-cost, objective tools; icterometers and smartphone apps with high diagnostic accuracy are promising alternatives.^42,43,46,56^ Second, prospective diagnostic accuracy studies of these tools against serum bilirubin are needed in community settings, with deliberate inclusion of preterm and dark-skinned infants. Third, because even standardized training did not overcome the poor sensitivity of the WHO IMCI method, the WHO should update its IMCI guidance to incorporate an objective measurement step rather than continuing to rely on visual inspection alone.

### Conclusion

In community settings across five low- and middle-income countries, the World Health Organization Integrated Management of Childhood Illnesses technique for visual inspection to identify jaundice showed very low sensitivity and positive percent agreement, with high consistent negative percent agreement, when compared with transcutaneous bilirubin measurement, indicating poor case detection but reliable exclusion of infants unlikely to require referral. Among preterm infants, sensitivity was modestly higher than in term infants but remained clinically inadequate, reinforcing that visual inspection alone is insufficient for this high-risk group. These findings indicate that the current WHO IMCI visual inspection method has limited utility and underscores the need for updated screening approaches in community settings where objective bilirubin measurement is unavailable.

## Data Availability

De-identified individual participant data may be made available to qualified researchers following review and approval by the PRISMA Consortium. Researchers interested in requesting access may contact the corresponding authors. Data sharing is subject to applicable ethical and regulatory requirements and execution of an appropriate data use agreement. The full study protocol and statistical analysis plan are available upon reasonable request. The published protocol paper is available here: https://bmjopen.bmj.com/content/16/1/e104512.long

## Acknowledgements

The authors are grateful to the participants who voluntarily took part in this study and the data collectors for their help in data collection. We appreciate the support of Dr Jacquline Asibey and Dr Rosemond Kokuro of the Holy Family Hospital in Techiman and St Theresa’s Hospital in Nkoranza respectively, in the training of study staff and healthcare providers in the study area in Ghana on IMCI. We would also like to thank additional members of the research teams at each study site, including: Harun Owuor from the Kenya Medical Research Institute, Kisumu, Kenya; Dr Arun Singh Jadaun, Dr Rupa Talukdar, Dr Kamal Kant, and Dr Meghna Singh from the Society of Applied Studies, New Delhi, India; Dr. Lankala Pramitha, Dr. Tobey Ann Marcus, and Lydia Vasanth from the Christian Medical College of Vellore, Vellore, India; Twaambo Munaumba, Emmanuel Mweni, Augustine Tunga, Caroline Mulenga, Sarah Mukuka, Inutu Matongo, Felistas Mbewe, and Rachael Ngulube from the University of North Carolina Global Projects Zambia, Lusaka, Zambia; and Charlotte Tawaiah from Kintampo Health Research Centre, Kintampo, Ghana. The Gates Foundation supported this work. The conclusions and opinions expressed in this work are, however, those of the author(s) alone and shall not be attributed to the Foundation. Under the grant conditions of the Foundation, a Creative Commons Attribution 4.0 License has already been assigned to the Author Accepted Manuscript version that might arise from this submission.

## PRISMA Consortium Collaborator Statement

The Pregnancy Risk, Infant Surveillance, and Measurement Alliance (PRISMA) Consortium includes the following institutions and members: **Aga Khan University** (Zahra Hoodbhoy, Fyezah Jehan, Amna Khan, Muhammad Imran Nisar, Asad Sheikh, Shayan Khakwani, Kinza Farooqui, Danish Hudani, Aamir Abbas, Muhammad Kashif, Nida Yazdani); **Beth Israel Deaconess Medical Center** (Blair J. Wylie); **Christian Medical College, Vellore** (Anne George Cherian, Santosh Joseph Benjamin, James A, Indhumathi, Daniel Jebakumar, Lydia Vasanth, Venkata Raghava Mohan, Vijayalekshmi B, Jayakumar Amirtharaj G, Pamela Christudas, John Jude Antony Prakash, Dhanalakshmi Solaimali, Rani Diana Sahni, John Fletcher, Asha Mary Abraham,Priya Abraham, Rajesh Kannangai, Divya M, Manish Kumar, Mintoo M Tergestina, Sebin George Abraham, Beena Koshy, Molly Jacob); **Dr. Dang’s Laboratory** (Leena Chatterjee, Arjun Dang, Manavi Dang, R Venketeshwar); **Gates Foundation** (Laura M. Lamberti); **The George Washington University Milken Institute School of Public Health** (Nazia Binte Ali, Sasha G. Baumann, Emma Cook, Bethany Freeman, Xinyi Li, Casey Kalman, Jaime Marquis, Jamie Minchin, Christopher N. Mores, Erin M. Oakley, Savannah O’Malley, Qing Pan, Abigale Proctor, Jennifer Seager, Alyssa Shapiro, Emily R. Smith, Ziwei Song, Precious Williams, Wen-Chien Yang); **Harvard T.H. Chan School of Public Health** (Christopher R. Sudfeld); **Kenya Medical Research Institute and Liverpool School of Tropical Medicine** (Victor Akelo, Florence Aweyo, Kephas Otieno, Harun Owuor, Dickens Onyango, Caleb Sagam, Feiko ter Kuile, Joyce Were, Zacchaeus Were, Dickson Gethi, Dorothy Lynda Achieng, Kevin Kasadhe, Edwin Kiplelgo); **Kintampo Health Research Centre** (Irene Apewe Adjei, Ken Ae-Ngibise, Veronica Agyemang, Japhet Anim, Kwaku Poku Asante, Ellen Boamah-Kaali, Richard Boakye, Stephaney Gyasse, Sam Newton, Eliezer Odei-Lartey, Samuel Addo Oppong, Richard Tetteh, Serwah Felicia, Solomon Nyame); **Society for Applied Studies** (Sarmila Mazumder, Neeraj Sharma, Arun Singh Jadaun, Rupa Talukdar, Mrinal Kishore, Dinesh Kumar Dhingra, Meghna Singh, Munita Jat, Kamal Kant, Soumya R. Nayak); University of Alabama at Birmingham School of Medicine (Lynda Ugwu); **University of North Carolina—Global Projects Zambia** (Margaret P. Kasaro, Felistas Mbewe, Twaambo Munaumba, Humphrey Mwape, Augustine Tunga); **University of North Carolina at Chapel Hill** (M. Bridget Spelke, Jeffrey S.A. Stringer); **University of Zambia** (Wilbroad Mutale, Mutale Sampa, Bellington Vwalika); and **VITAL Pakistan Trust** (Kinza Farooqui, Farzana Shaheen).

## Funding

No funding was received for this secondary data analysis. The primary pregnancy cohort was funded by the Bill and Melinda Gates Foundation, grant numbers (INV-047400 to KPA and SN; INV-057219 to VA; INV-043092 to SB; INV-057220 to ZH; INV-016221 to MPK; INV-057222 to WM; INV-041999 to ERS; INV-060797 to CNM; and INV-057223 to SM).

## Author contributions

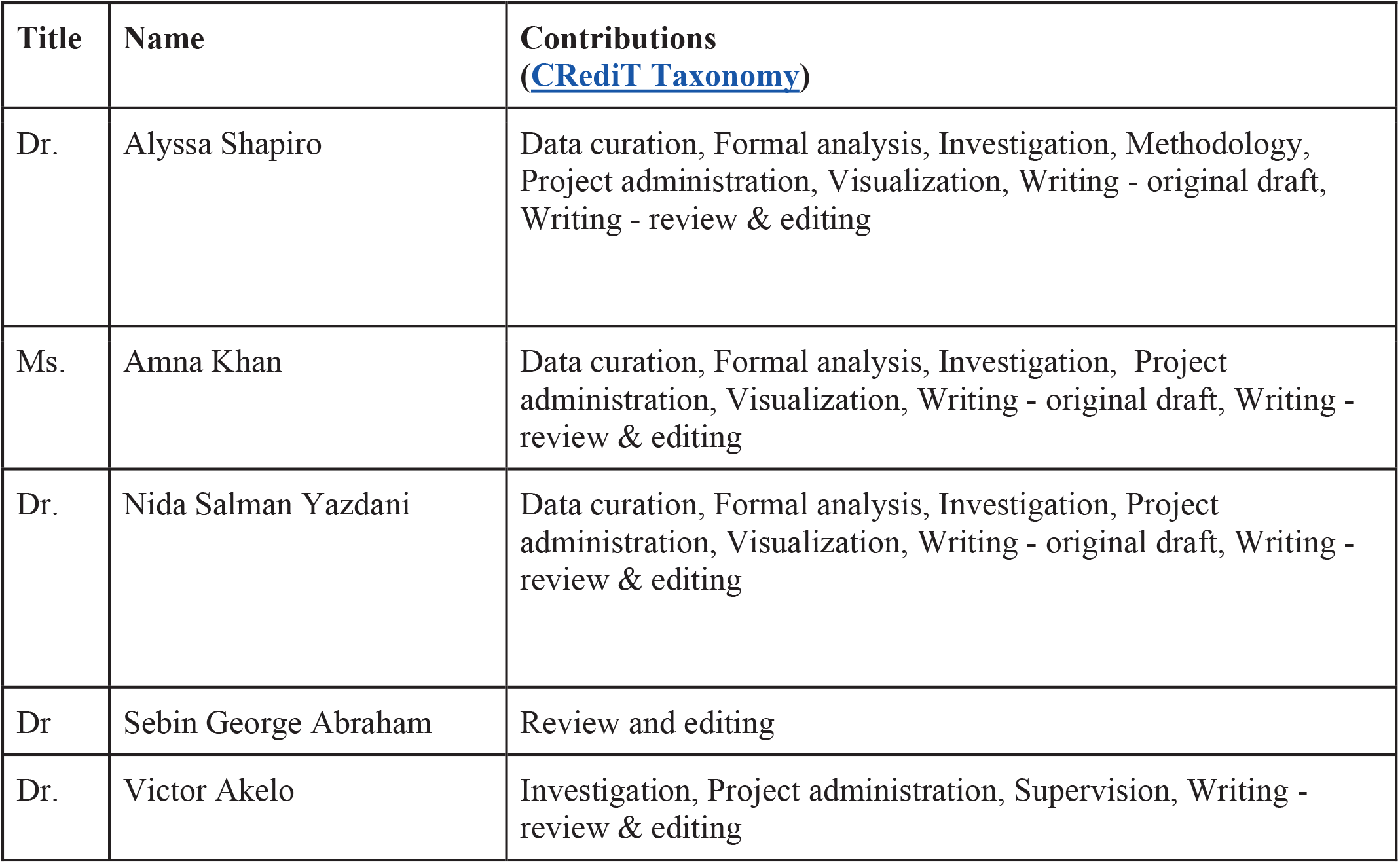
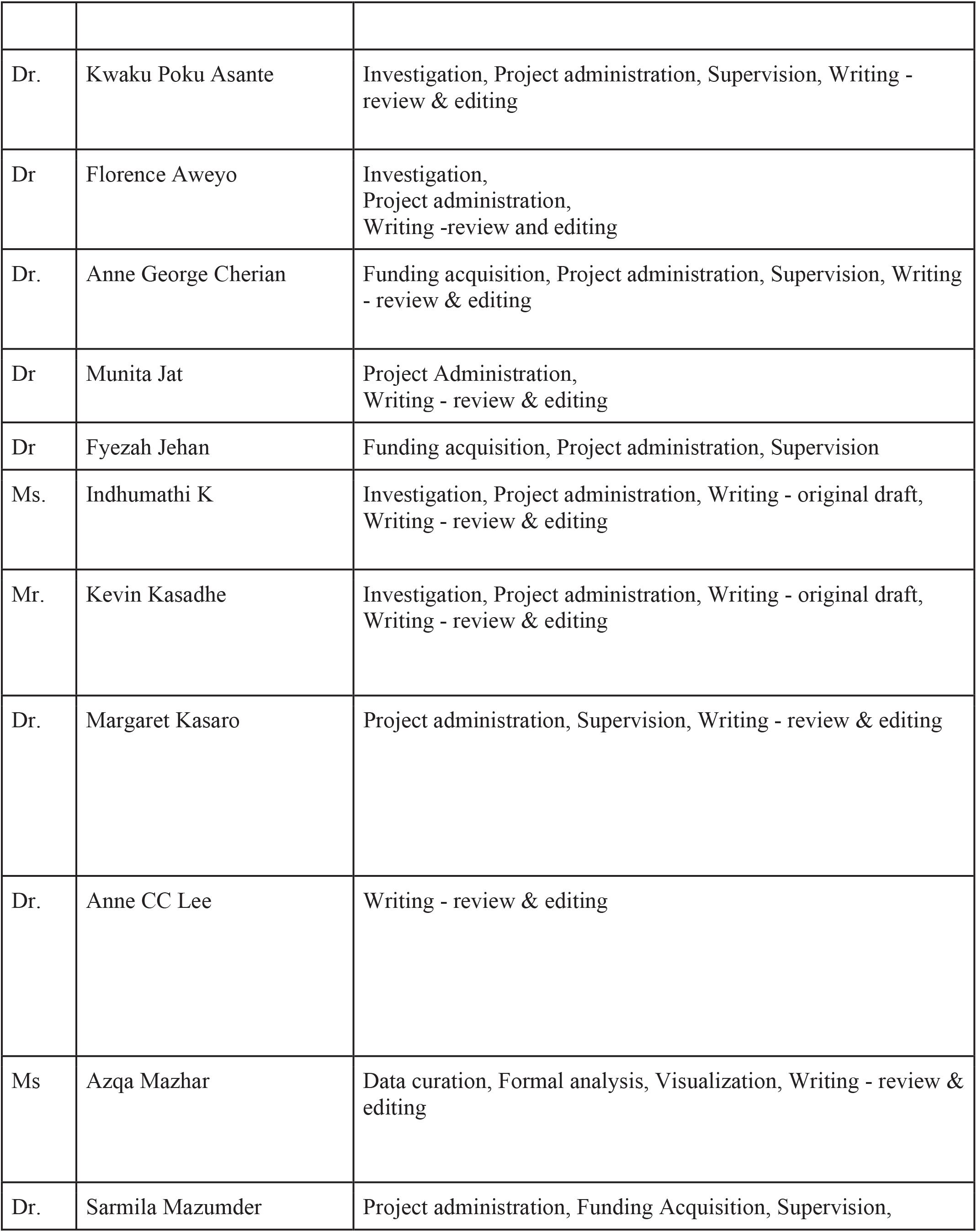
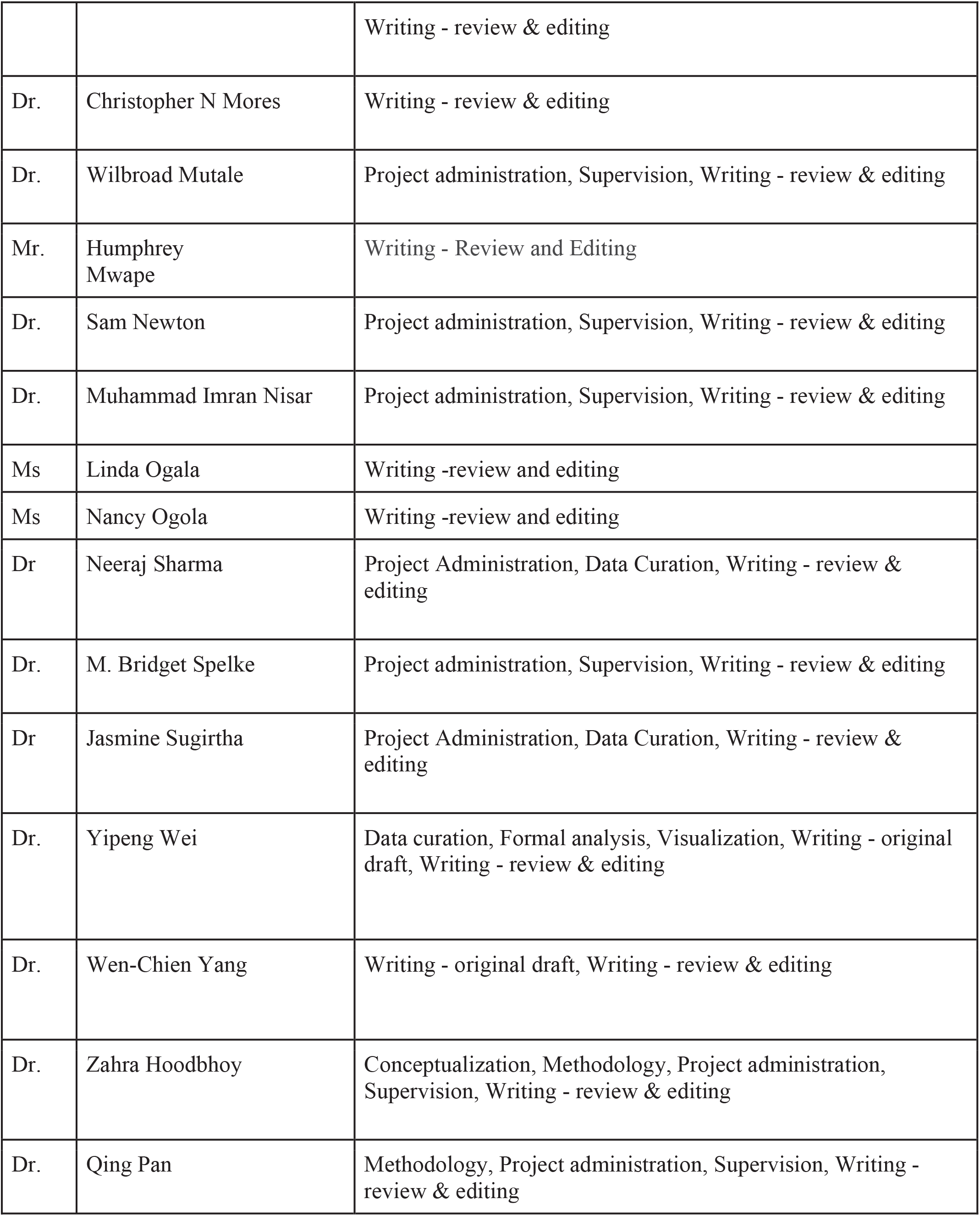
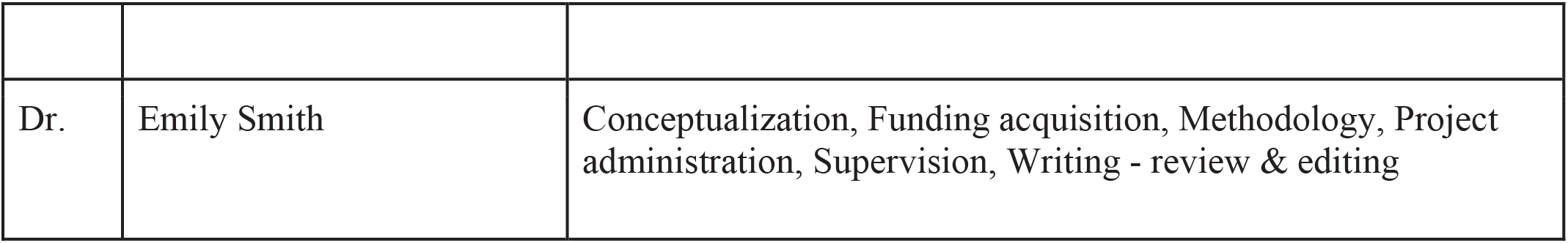

## Ethics declarations

### Consent statement

Informed consent was obtained from participants of the PRISMA MNH study (who are the parents of all study participants in this work) at the time of original data collection.

### Competing interests

**Dr. Alyssa Shapiro** is an inventor of the BiliDx which has been licensed to 3rd Stone Design; the devices are licensed at 0% royalty in GAVI-eligible countries and the inventor’s share of all royalties has been donated to Rice University. Dr. Shapiro has also received grant funding from the Thrasher Research Fund for research on the Bili-ruler.

**Dr. CC Lee** invented the Biliruler and was previously employed by Brigham and Women’s Hospital, who granted a sole, royalty-bearing license to Little Sparrows Technologies under the copyright rights to use, reproduce, and sell licensed products in high-income countries. Dr. Lee has received grant funding from Saving Lives at Birth for research on the Bili-ruler.

**Dr. Wilbroad Mutale** is the business partner for Picterus in Africa.

These declarations do not alter the authors’ adherence to all policies on sharing data and materials

### Ethical approval

The PRISMA MNH study was approved by the George Washington University’s Committee on Human Research (IRB: FWA00005945) on September 30, 2022 and received local and national ethical approval in Pakistan (Aga Khan University ERC 2022-5920-22763, and Pakistan National Bioethics Committee 4-87/NBC-58/8/22/337), Kenya (KEMRI Scientific and Ethics Review Unit KEMRI/SERU/CGHR/04/10/358/4166; Liverpool 23-020), Zambia (University of Zambia Biomedical Research Ethics Committee: 016-04-14 and University of North Carolina Chapel Hill Office of Human Research Ethics: 356795), Ghana (Kintampo Health Research Centre Institutional Ethics Committee (IEC) FWA00011103; Ref 0004854 and Ghana Health Service Ethics Review Committee FWA00020025), Vellore, India (Christian Medical College Vellore Office of Research IRB No 14553), and Hodal, India (Ethics Review Committee, Society for Applied Studies, SAS/ERC/ReMAPP Study/2022). Work in this manuscript is covered by the IRB and informed consents of the overall PRISMA MNH study.

## Notes

### Clinical Protocols

https://bmjopen.bmj.com/content/16/1/e104512.long

### Author Declarations

The PRISMA MNH study was approved by the George Washington University Committee on Human Research (IRB: FWA00005945) on September 30, 2022 and received local and national ethical approval in Pakistan (Aga Khan University Ethics Review Committee (ERC) 2022-5920-22763 and Pakistan National Bioethics Committee 4-87/NBC-58/8/22/337), Kenya (KEMRI Scientific and Ethics Review Unit KEMRI/SERU/CGHR/04/10/358/4166; Liverpool 23-020), Zambia (University of Zambia Biomedical Research Ethics Committee: 016-04-14 and University of North Carolina Chapel Hill Office of Human Research Ethics: 356795), Ghana (Kintampo Health Research Centre Institutional Ethics Committee (IEC) FWA00011103; Ref 0004854 and Ghana Health Service Ethics Review Committee FWA00020025), Vellore, India (Christian Medical College Vellore Office of Research IRB No 14553), and Hodal, India (Ethics Review Committee, Society for Applied Studies, SAS/ERC/ReMAPP Study/2022). Work in this manuscript is covered by the IRB and informed consents of the overall PRISMA MNH study.

