## Supplementary Information for "Diagnostic Accuracy of WHO IMCI Visual Inspection for Neonatal Jaundice: A Prospective Multi-Country Cohort"

### Supplemental Information

**Table S1.** STARD 2015 checklist

| Section & Topic | No | Item | Reported on page # |
|---|---|---|---|
| **TITLE OR ABSTRACT** | | | |
| | **1** | Identification as a study of diagnostic accuracy using at least one measure of accuracy<br>(such as sensitivity, specificity, predictive values, or AUC) | Cover page (title) |
| **ABSTRACT** | | | |
| | **2** | Structured summary of study design, methods, results, and conclusions<br>(for specific guidance, see STARD for Abstracts) | Cover page (abstract) |
| **INTRODUCTION** | | | |
| | **3** | Scientific and clinical background, including the intended use and clinical role of the index test | 1-3 |
| | **4** | Study objectives and hypotheses | 3 |
| **METHODS** | | | |
| *Study design* | **5** | Whether data collection was planned before the index test and reference standard<br>were performed (prospective study) or after (retrospective study) | 3 |
| *Participants* | **6** | Eligibility criteria | 3-4 |
| | **7** | On what basis potentially eligible participants were identified<br>(such as symptoms, results from previous tests, inclusion in registry) | 3-4 |
| | **8** | Where and when potentially eligible participants were identified (setting, location and dates) | 3-4 |
| | **9** | Whether participants formed a consecutive, random or convenience series | 3 |
| *Test methods* | **10a** | Index test, in sufficient detail to allow replication | 4-5 |
| | **10b** | Reference standard, in sufficient detail to allow replication | 5 |
| | **11** | Rationale for choosing the reference standard (if alternatives exist) | 5-6 |
| | **12a** | Definition of and rationale for test positivity cut-offs or result categories<br>of the index test, distinguishing pre-specified from exploratory | 5 |
| | **12b** | Definition of and rationale for test positivity cut-offs or result categories<br>of the reference standard, distinguishing pre-specified from exploratory | 6-7 |
| | **13a** | Whether clinical information and reference standard results were available<br>to the performers/readers of the index test | 5 |
| | **13b** | Whether clinical information and index test results were available<br>to the assessors of the reference standard | 5 |
| *Analysis* | **14** | Methods for estimating or comparing measures of diagnostic accuracy | 7 |
| | **15** | How indeterminate index test or reference standard results were handled | 7 |
| | **16** | How missing data on the index test and reference standard were handled | 7 |
| | **17** | Any analyses of variability in diagnostic accuracy, distinguishing pre-specified from exploratory | 7 |
| | **18** | Intended sample size and how it was determined | 3 |
| **RESULTS** | | | |
| *Participants* | **19** | Flow of participants, using a diagram | 8 (Figure 1) |
| | **20** | Baseline demographic and clinical characteristics of participants | 8 (Table 1) |
| | **21a** | Distribution of severity of disease in those with the target condition | 8 (Table S2) |
| | **21b** | Distribution of alternative diagnoses in those without the target condition | 8 (Table S2) |
| | **22** | Time interval and any clinical interventions between index test and reference standard | 5 |
| *Test results* | **23** | Cross tabulation of the index test results (or their distribution)<br>by the results of the reference standard | 8 (Table S2) |
| | **24** | Estimates of diagnostic accuracy and their precision (such as 95% confidence intervals) | 8-9 (Figure 2) |
| | **25** | Any adverse events from performing the index test or the reference standard | 8 |
| **DISCUSSION** | | | |

CC BY SA

| | | | |
|---|---|---|---|
| | **26** | Study limitations, including sources of potential bias, statistical uncertainty, and generalisability | 14-15 |
| | **27** | Implications for practice, including the intended use and clinical role of the index test | 15-16 |
| **OTHER INFORMATION** | | | |
| | **28** | Registration number and name of registry | 3 |
| | **29** | Where the full study protocol can be accessed | 17 |
| | **30** | Sources of funding and other support; role of funders | 21 |

**Table S2.** Prevalence of neonatal jaundice, assessed by visual inspection and transcutaneous bilirubinometry, overall and stratified by study site

| Characteristic | Overall | North India | South India | Pakistan | Ghana | Kenya | Zambia |
|---|---|---|---|---|---|---|---|
| Number of infants represented in the study | 14,331 | 1,798 | 2,053 | 4,780 | 2,036 | 2,275 | 1,389 |
| **Visit 1 (0-3 Days)** | | | | | | | |
| N = | 10,613 | 1,332 | 1,807 | 3,846 | 858 | 1,623 | 1,147 |
| **Visual Inspection** | | | | | | | |
| Denominator (valid VI result) | 10,613 | 1,332 | 1,807 | 3,846 | 858 | 1,623 | 1,147 |
| Jaundiced | 102 (1.0%) | 10 (0.8%) | 3 (0.2%) | 62 (1.6%) | 11 (1.3%) | 8 (0.5%) | 8 (0.7%) |
| Severely Jaundiced | 272 (2.6%) | 31 (2.3%) | 10 (0.6%) | 181 (4.7%) | 15 (1.7%) | 33 (2.0%) | 2 (0.2%) |
| **TCB** | | | | | | | |
| *TCB ≥ 15* | | | | | | | |
| Denominator (valid TCB)[1] | 10,613 | 1,332 | 1,807 | 3,846 | 858 | 1,623 | 1,147 |
| N (%) | 204 (1.9%) | 37 (2.8%) | 9 (0.5%) | 70 (1.8%) | 35 (4.1%) | 39 (2.4%) | 14 (1.2%) |
| *TCB ≥ NICE Threshold* | | | | | | | |
| Denominator (valid NICE threshold)[2] | 10,613 | 1,332 | 1,807 | 3,846 | 858 | 1,623 | 1,147 |
| N (%) | 6,108 (58%) | 699 (52%) | 1,188 (66%) | 1,915 (50%) | 508 (59%) | 1,089 (67%) | 709 (62%) |
| *TCB ≥ AAP Threshold* | | | | | | | |
| Denominator (valid AAP threshold)[3] | 10,289 | 1,309 | 1,779 | 3,684 | 832 | 1,582 | 1,103 |
| N (%) | 2,668 (26%) | 334 (26%) | 469 (26%) | 677 (18%) | 262 (31%) | 641 (41%) | 285 (26%) |
| **Visit 2 (3-5 Days)** | | | | | | | |
| N = | 7,255 | 1,157 | 893 | 2,295 | 1,003 | 1,356 | 551 |
| **Visual Inspection** | | | | | | | |
| Denominator (valid VI result) | 7,255 | 1,157 | 893 | 2,295 | 1,003 | 1,356 | 551 |
| Jaundiced | 992 (14%) | 83 (7.2%) | 120 (13%) | 646 (28%) | 25 (2.5%) | 86 (6.3%) | 32 (5.8%) |
| Severely Jaundiced | 309 (4.3%) | 47 (4.1%) | 61 (6.8%) | 118 (5.1%) | 28 (2.8%) | 34 (2.5%) | 21 (3.8%) |
| **TCB** | | | | | | | |
| *TCB ≥ 15* | | | | | | | |
| Denominator (valid TCB)[1] | 7,244 | 1,157 | 889 | 2,295 | 1,003 | 1,349 | 551 |
| N (%) | 824 (11%) | 145 (13%) | 114 (13%) | 256 (11%) | 112 (11%) | 131 (9.7%) | 66 (12%) |
| *TCB ≥ NICE Threshold* | | | | | | | |
| Denominator (valid NICE threshold)[2] | 7,244 | 1,157 | 889 | 2,295 | 1,003 | 1,349 | 551 |

| | | | | | | | |
|---|---|---|---|---|---|---|---|
| N (%) | 1,113 (15%) | 203 (18%) | 188 (21%) | 402 (18%) | 94 (9.4%) | 151 (11%) | 75 (14%) |
| *TCB ≥ AAP Threshold* | | | | | | | |
| Denominator (valid AAP threshold)[3] | 7,019 | 1,131 | 868 | 2,192 | 982 | 1,310 | 536 |
| N (%) | 430 (6.1%) | 62 (5.5%) | 86 (9.9%) | 134 (6.1%) | 51 (5.2%) | 64 (4.9%) | 33 (6.2%) |
| **Visit 3 (5-14 Days)** | | | | | | | |
| N = | 12,756 | 1,748 | 1,738 | 4,242 | 1,703 | 2,112 | 1,213 |
| **Visual Inspection** | | | | | | | |
| Denominator (valid VI result) | 12,756 | 1,748 | 1,738 | 4,242 | 1,703 | 2,112 | 1,213 |
| Jaundiced | 1,389 (11%) | 100 (5.7%) | 54 (3.1%) | 1,046 (25%) | 43 (2.5%) | 116 (5.5%) | 30 (2.5%) |
| Severely Jaundiced | 281 (2.2%) | 51 (2.9%) | 35 (2.0%) | 143 (3.4%) | 6 (0.4%) | 31 (1.5%) | 15 (1.2%) |
| **TCB** | | | | | | | |
| *TCB ≥ 15* | | | | | | | |
| Denominator (valid TCB)[1] | 12,751 | 1,745 | 1,738 | 4,242 | 1,703 | 2,110 | 1,213 |
| N (%) | 698 (5.5%) | 140 (8.0%) | 23 (1.3%) | 347 (8.2%) | 51 (3.0%) | 78 (3.7%) | 59 (4.9%) |
| *TCB ≥ NICE Threshold* | | | | | | | |
| Denominator (valid NICE threshold)[2] | 12,751 | 1,745 | 1,738 | 4,242 | 1,703 | 2,110 | 1,213 |
| N (%) | 1,086 (8.5%) | 193 (11%) | 85 (4.9%) | 569 (13%) | 59 (3.5%) | 107 (5.1%) | 73 (6.0%) |
| *TCB ≥ AAP Threshold* | | | | | | | |
| Denominator (valid AAP threshold)[3] | 12,333 | 1,698 | 1,706 | 4,037 | 1,673 | 2,049 | 1,170 |
| N (%) | 257 (2.1%) | 47 (2.8%) | 9 (0.5%) | 138 (3.4%) | 18 (1.1%) | 29 (1.4%) | 16 (1.4%) |

[1]TCB ≥ 15 is recommended by the AAP as a criteria for defining newborns at risk of follow-up care.

[2]NICE guidelines contain nomograms which illustrate cutoff values based on gestational age, neonatal age, and risk factors. For each assessment, the NICE cutoff value was determined from the nomograms, and the newborn was determined to be 'Positive' or 'Negative' or NNJ based on this cutoff value. If the gestational age at birth was < 23 weeks, the NICE cutoff value for 23 weeks was used, because the NICE guidelines do not give clear guidelines for infants with gestational age below 23 weeks.

[3]AAP guidelines contain nomograms which illustrate cutoff values based on gestational age, neonatal age, and risk factors. For each assessment, the AAP cutoff value was determined from the nomograms, and the newborn was determined to be 'Positive' or 'Negative' or NNJ based on this cutoff value. If the gestational age at birth was < 35 weeks, the infant was excluded from analysis, because the AAP guidelines do not give clear guidelines for infants in this category.

**Figure S1.** Diagnostic performance of visual inspection against transcutaneous bilirubin, using alternate methods of defining "Positive for Jaundice" by transcutaneous bilirubin

Legend (%): 100, 75, 50, 25

a)

| Sensitivity (Se) | | | |
|---|---|---|---|
| Site | 0-3 Days | 3-5 Days | 5-14 Days |
| Pooled | 25 | 19 | 13 |
| North India | 25 | 21 | 24 |
| South India | 38 | 29 | 30 |
| Pakistan | 40 | 15 | 9 |
| Ghana | 12 | 10 | 10 |
| Kenya | 14 | 20 | 6 |
| Zambia | 10 | 22 | 12 |

| Specificity (Sp) | | | |
|---|---|---|---|
| Site | 0-3 Days | 3-5 Days | 5-14 Days |
| Pooled | 98 | 98 | 99 |
| North India | 98 | 98 | 99 |
| South India | 100 | 97 | 99 |
| Pakistan | 96 | 97 | 98 |
| Ghana | 99 | 98 | 100 |
| Kenya | 98 | 99 | 99 |
| Zambia | 100 | 98 | 100 |

| Positive Percent Agreement (PPA) | | | |
|---|---|---|---|
| Site | 0-3 Days | 3-5 Days | 5-14 Days |
| Pooled | 20 | 53 | 31 |
| North India | 32 | 64 | 63 |
| South India | 33 | 58 | 29 |
| Pakistan | 16 | 35 | 17 |
| Ghana | 27 | 42 | 67 |
| Kenya | 17 | 72 | 17 |
| Zambia | 50 | 63 | 56 |

| Negative Percent Agreement (NPA) | | | |
|---|---|---|---|
| Site | 0-3 Days | 3-5 Days | 5-14 Days |
| Pooled | 99 | 91 | 97 |
| North India | 98 | 91 | 95 |
| South India | 100 | 90 | 99 |
| Pakistan | 99 | 92 | 96 |
| Ghana | 96 | 90 | 98 |
| Kenya | 98 | 93 | 97 |
| Zambia | 99 | 91 | 97 |

b)

| Sensitivity (Se) | | | |
|---|---|---|---|
| Site | 0-3 Days | 3-5 Days | 5-14 Days |
| Pooled | 18 | 11 | 9 |
| North India | 4 | 7 | 9 |
| South India | 6 | 13 | 15 |
| Pakistan | 29 | 15 | 9 |
| Ghana | 0 | 10 | 0 |
| Kenya | 3 | 3 | 9 |
| Zambia | 0 | 6 | 10 |

| Specificity (Sp) | | | |
|---|---|---|---|
| Site | 0-3 Days | 3-5 Days | 5-14 Days |
| Pooled | 99 | 98 | 98 |
| North India | 99 | 100 | 99 |
| South India | 100 | 90 | 95 |
| Pakistan | 99 | 98 | 97 |
| Ghana | 100 | 100 | 100 |
| Kenya | 98 | 100 | 100 |
| Zambia | 100 | 100 | 100 |

| Positive Percent Agreement (PPA) | | | |
|---|---|---|---|
| Site | 0-3 Days | 3-5 Days | 5-14 Days |
| Pooled | 89 | 93 | 87 |
| North India | 67 | 100 | 94 |
| South India | 100 | 75 | 79 |
| Pakistan | 94 | 94 | 84 |
| Ghana | NA | 100 | NA |
| Kenya | 33 | 100 | 100 |
| Zambia | NA | 100 | 100 |

| Negative Percent Agreement (NPA) | | | |
|---|---|---|---|
| Site | 0-3 Days | 3-5 Days | 5-14 Days |
| Pooled | 82 | 35 | 42 |
| North India | 69 | 21 | 32 |
| South India | 88 | 32 | 50 |
| Pakistan | 83 | 39 | 39 |
| Ghana | 82 | 35 | 51 |
| Kenya | 74 | 38 | 54 |
| Zambia | 90 | 43 | 54 |

c)

| Sensitivity (Se) | | | |
|---|---|---|---|
| Site | 0-3 Days | 3-5 Days | 5-14 Days |
| Pooled | 47 | 21 | 14 |
| North India | 100 | 21 | 25 |
| South India | 100 | 14 | 33 |
| Pakistan | 67 | 25 | 10 |
| Ghana | 0 | 8 | 0 |
| Kenya | 0 | 11 | 31 |
| Zambia | 0 | 18 | 21 |

| Specificity (Sp) | | | |
|---|---|---|---|
| Site | 0-3 Days | 3-5 Days | 5-14 Days |
| Pooled | 97 | 95 | 96 |
| North India | 99 | 98 | 98 |
| South India | 100 | 88 | 90 |
| Pakistan | 95 | 93 | 95 |
| Ghana | 100 | 93 | 100 |
| Kenya | 98 | 100 | 99 |
| Zambia | 100 | 100 | 98 |

| Positive Percent Agreement (PPA) | | | |
|---|---|---|---|
| Site | 0-3 Days | 3-5 Days | 5-14 Days |
| Pooled | 25 | 49 | 42 |
| North India | 33 | 75 | 75 |
| South India | 100 | 12 | 7 |
| Pakistan | 25 | 49 | 35 |
| Ghana | NA | 25 | NA |
| Kenya | 0 | 100 | 71 |
| Zambia | NA | 100 | 67 |

| Negative Percent Agreement (NPA) | | | |
|---|---|---|---|
| Site | 0-3 Days | 3-5 Days | 5-14 Days |
| Pooled | 99 | 83 | 84 |
| North India | 100 | 84 | 84 |
| South India | 100 | 89 | 98 |
| Pakistan | 99 | 82 | 79 |
| Ghana | 96 | 78 | 89 |
| Kenya | 97 | 84 | 93 |
| Zambia | 97 | 84 | 88 |

d)

| Sensitivity (Se) | | | |
|---|---|---|---|
| Site | 0-3 Days | 3-5 Days | 5-14 Days |
| Pooled | 28 | 20 | 14 |
| North India | 27 | 21 | 24 |
| South India | 44 | 28 | 30 |
| Pakistan | 47 | 19 | 10 |
| Ghana | 11 | 10 | 8 |
| Kenya | 13 | 19 | 12 |
| Zambia | 7 | 21 | 15 |

| Specificity (Sp) | | | |
|---|---|---|---|
| Site | 0-3 Days | 3-5 Days | 5-14 Days |
| Pooled | 98 | 98 | 98 |
| North India | 98 | 98 | 99 |
| South India | 100 | 96 | 98 |
| Pakistan | 96 | 97 | 97 |
| Ghana | 99 | 98 | 100 |
| Kenya | 98 | 99 | 99 |
| Zambia | 100 | 99 | 99 |

| Positive Percent Agreement (PPA) | | | |
|---|---|---|---|
| Site | 0-3 Days | 3-5 Days | 5-14 Days |
| Pooled | 21 | 52 | 35 |
| North India | 32 | 66 | 67 |
| South India | 40 | 52 | 20 |
| Pakistan | 18 | 41 | 24 |
| Ghana | 27 | 39 | 67 |
| Kenya | 15 | 74 | 29 |
| Zambia | 50 | 67 | 60 |

| Negative Percent Agreement (NPA) | | | |
|---|---|---|---|
| Site | 0-3 Days | 3-5 Days | 5-14 Days |
| Pooled | 99 | 90 | 95 |
| North India | 98 | 90 | 94 |
| South India | 100 | 90 | 99 |
| Pakistan | 99 | 90 | 92 |
| Ghana | 96 | 90 | 97 |
| Kenya | 98 | 92 | 97 |
| Zambia | 99 | 90 | 96 |
